# Molecular tracing of SARS-CoV-2 in Italy in the first three months of the epidemic

**DOI:** 10.1101/2020.07.06.20147140

**Authors:** Alessia Lai, Annalisa Bergna, Sara Caucci, Nicola Clementi, Ilaria Vicenti, Filippo Dragoni, Anna Maria Cattelan, Stefano Menzo, Angelo Pan, Annapaola Callegaro, Adriano Tagliabracci, Arnaldo Caruso, Francesca Caccuri, Silvia Ronchiadin, Claudia Balotta, Maurizio Zazzi, Emanuela Vaccher, Massimo Clementi, Massimo Galli, Gianguglielmo Zehender, on behalf of SARS-CoV-2 ITALIAN RESEARCH ENTERPRISE – (SCIRE) collaborative Group

## Abstract

The aim of this study is the characterization and genomic tracing by phylogenetic analyses of 59 new SARS-CoV-2 Italian isolates obtained from patients attending clinical centres in North and Central Italy until the end of April 2020.

All but one of the newly characterized genomes belonged to the lineage B.1, the most frequently identified in European countries, including Italy. Only a single sequence was found to belong to lineage B.

A mean of 6 nucleotide substitutions per viral genome was observed, without significant differences between synonymous and non-synonymous mutations, indicating genetic drift as a major source for virus evolution.

tMRCA estimation confirmed the probable origin of the epidemic between the end of January and the beginning of February with a rapid increase in the number of infections between the end of February and mid-March. Since early February, an effective reproduction number (R_e_) greater than 1 was estimated, which then increased reaching the peak of 2.3 in early March, confirming the circulation of the virus before the first COVID-19 cases were documented.

Continuous use of state-of-the-art methods for molecular surveillance is warranted to trace virus circulation and evolution and inform effective prevention and containment of future SARS-CoV-2 outbreaks.

## 1. Introduction

Italy is one of the countries most and earlier affected in Europe by the COVID-19 pandemic (https://gisanddata.maps.arcgis.com/apps/opsdashboard/index.html#/bda7594740fd40299423467b48e9ecf6). The first autochthonous cases of Coronavirus 2019 Disease (COVID-19) were observed starting from February 21, 2020 in Codogno (Lodi province), determining on February 22, 2020 the establishment of a ‘red zone’ to contain the epidemic, encompassing 11 municipalities. Thereafter, in a short time, it became evident that the epidemic had already involved a large part of Lombardy region and then spread to neighbouring regions and, substantially less, to the rest of the country. On March 9, lockdown was declared for the entire country. The rapidly increasing number of patients who required hospitalization in the intensive care unit suggested that the virus may have circulated for a long period and caused thousands of contagions before the epidemic became manifest [1].

SARS-CoV-2 was first detected in Italy in a couple of Chinese tourists coming from Wuhan on January 31 [2]. Subsequent evaluations have not shown a relationship between the sequence of these strains and those implicated in the epidemic in Lombardy [3].

On the contrary, the Codogno strains resulted strictly related with a strain of SARS-CoV-2 coming from Shanghai which caused a small outbreak in Munich around January 20 [1] and was probably spread later to other European countries and beyond the Atlantic [4]. These sequences are part of a clade initially defined as a European clade, the old Nexstrain A2a subclade, which is currently the most widespread outside China and probably responsible for most of the world pandemic [5].

In the face of more than 240,000 notified cases in Italy, the entire genomes available in public databases are still scarce (77 at the time of this study). The availability of large numbers of sequences collected over time is necessary for molecular surveillance of the epidemic and for evaluation and planning of effective control strategies. To perform this study, a network of Italian Clinical centres and Laboratories across Italy generated additional 59 full-length SARS-CoV-2 sequences from COVID-19 patients ranging from the end of February to the end of April. This contribution helps to trace the temporal origin, the rate of viral evolution and the population dynamics of SARS-CoV-2 in Italy by phylogeny.

## 2. Materials and Methods

### 2.1 Patients and Methods

A total of 59 SARS-CoV-2 whole genomes were newly characterized from an equal number of patients affected by COVID-19, attending different clinical centres in Northern and Central Italy, from the beginning of the epidemic (February 22, 2020) until April 27, 2020 (Table S1).

All of the data used in this study were previously anonymised as required by the Italian Data Protection Code (Legislative Decree 196/2003) and the general authorisations issued by the Data Protection Authority. Ethics Committee approval was deemed unnecessary because, under Italian law, all sensitive data were deleted and we collected only age, gender and sampling date (Art. 6 and Art. 9 of Legislative Decree 211/2003).

Eighteen sequences were obtained after isolating the virus in Vero E6 cells while the remaining 41 were obtained directly from biological samples such as nasopharyngeal swabs or broncho-alveolar lavages (39 and 2, respectively).

SARS-CoV-2 RNA was extracted using the Kit QIAsymphony DSP Virus/Pathogen Midi kit on the QIAsymphony automated platform (QIAGEN, Hilden, Germany) (n=9) and manually with QIAamp Viral RNA Mini Kit (n=50).

Full genome sequences were obtained with different protocols by amplifying 26 fragments as previously described (n=42) [1] or using random hexamer primers (n=8) or Ion AmpliSeq SARS-CoV-2 Research Panel (Thermo Fisher Scientific) (n=9). The PCR products were used to prepare a library for Illumina deep sequencing using a Nextera XT DNA Sample Preparation and Index kit (Illumina, San Diego, California, USA) in accordance with the manufacturer’s manual, and sequencing was carried out on a Illumina MiSeq platform for fifty samples, while the remaining nine were sequenced on Ion GeneStudio™ S5 System (Thermo Fisher Scientific) instrument following the Ion AmpliSeq™ RNA libraries protocol. The results were mapped and aligned to the reference genome obtained from GISAID (https://www.gisaid.org/, accession ID: EPI_ISL_412973) using Geneious software, v. 9.1.5 (http://www.geneious.com) [6] or Torrent Suite v. 5.10.1 or BWA-mem and rescued using Samtools alignment/Map (v 1.9).

### 2.2 Sequence data sets

The newly characterized 59 genomes plus three previously characterized isolates by us (EPI_ISL_417445-417447) [1] were aligned with a total of 77 Italian sequences available in public databases (GISAID, https://www.gisaid.org/) on May 13, 2020 and 452 genomes sampled in different European and Asian countries (513 and 16, respectively) representing all the different viral clades described in the Nextstrain platform (https://nextstrain.org/). The final data set thus included 588 sequences. Due to the large amount of available sequences, we focused the analysis on European strains by randomly selecting sequences from each country and by excluding identical strains or strains with more than 5% of gaps. We sampled the data in order to have no temporal gaps, by grouping the sequences by country/week/clade and randomly selecting the sequences in each group. We choose 15 sequences for clade A2 and 5 sequences for other clades for each European country. For countries with less than the required sequence number we kept all the sequences. The sampling dates of the entire dataset ranged from December 30, 2019 to April 27, 2020. Table S2 shows the accession IDs, sampling dates and locations of the sequences included in the dataset.

A subset of sequences assigned to the old Nextstrain A2 clade was generated for dating the epidemic, including all the Italian sequences, one German (EPI_ISL_406862) and three Chinese isolates from Shanghai, ancestral to the A2 clade (EPI_ISL_416327, EPI_ISL_416334 and EPI_ISL_416386). Coalescent and birth-death phylodynamic analyses were performed on the 136 Italian A2 sequences only.

Alignment was performed using MAFFT [7] and manually cropped to a final length of 29,779 bp using BioEdit v. 7.2.6.1 (http://www.mbio.ncsu.edu/bioedit/bioedit.html).

### 2.3 Genetic distance, recombination and selection pressure analyses

The MEGA X program was used to evaluate the genetic distance between and within Italian sequences on the full length genome, with variance estimation performed using 1,000 bootstrap replicates [8].

The RDP5 software was used to investigate the presence of potential recombination [9].

All of the genes were tested for selection pressure using Datamonkey (https://www.datamonkey.org/).

### 2.4 Phylogenetic and phylodynamic analyses

The simplest evolutionary model best fitting the sequence data was selected using the JmodelTest v.2.1.7 software [10], and proved to be the Hasegawa-Kishino-Yano model with a proportion of invariant sites (HKY+I).

The phylogenetic analysis for clade assignment was performed by RaxML [11] on the entire dataset of 588 genomes. During the period in which we were carrying out the study, the SARS-CoV-2 clade nomenclature system changed. In particular, Rambaut et al. proposed a dynamic nomenclature based on phylogenetic lineages, called Pangolin (Phylogenetic Assignment of Named Global Outbreak LINeages) [12]. For this reason we used the old Nextstrain and the new Pangolin (freely available at https://pangolin.cog-uk.io/) systems for strain classification. The new Nextstrain classification was performed by using the available script (https://github.com/nextstrain/ncov/blob/master/docs/running.md).

The virus’ phylogeny, evolutionary rates, times of the most recent common ancestor (tMRCA) and demographic growth were co-estimated in a Bayesian framework using a Markov Chain Monte Carlo (MCMC) method implemented in v.1.10.4 and v.2.62 of the BEAST package [13], [14].

A root-to-tip regression analysis was made using TempEst in order to investigate the temporal signal of the dataset [15].

Different coalescent priors (constant population size and exponential growth and Bayesian skyline) and strict *vs*. relaxed molecular clock models were tested by means of Path Sampling (PS) and Stepping Stone (SS) sampling [16]. The evolutionary rate prior normal distribution, after informing the mean evolutionary rate, was set at mean 0.8 × 10^−3^ substitutions/site/year (http://virological.org/t/phylodynamic-analysis-176-genomes-6-mar-2020/356).

The MCMC analysis was run until convergence with sampling every 10,000 generations. Convergence was assessed by estimating the effective sampling size (ESS) after 10% burn-in using Tracer v.1.7 software (http://tree.bio.ed.ac.uk/software/tracer/), and accepting ESS values of 200 or more. The uncertainty of the estimates was indicated by 95% highest marginal likelihoods estimated [17] by path sampling/stepping stone methods [16].

The final trees were summarised by selecting the tree with the maximum product of posterior probabilities (pp) (maximum clade credibility or MCC) after a 10% burn-in using Tree Annotator v.1.10.4 (included in the BEAST package), and were visualised using FigTree v.1.4.2 (http://tree.bio.ed.ac.uk/software/figtree/).

### 2.5 Birth-Death Skyline estimates of the effective reproductive number (R_e_)

The birth-death skyline model implemented in Beast 2.62 was used to infer changes in the effective reproductive number (R_e_), and other epidemiological parameters such as the death/recovery rate (δ), the transmission rate (λ), the origin of the epidemic, and the sampling proportion (ρ) [18]. Given that the samples were collected during a short period of time, a “birth-death contemporary” model was used.

The analyses were based on the previously selected HKY substitution model and the evolutionary rate was set to the value of 0.8 × 10^−3^ subs/site/year, which corresponds to the mean substitution rate estimated using a relaxed clock under the exponential coalescent model as transformed into units per year.

For the birth-death skyline analysis, from one to two R_e_ intervals and a log-normal prior with a mean (M) of 0.0 and a variance (S) of 1.0 were chosen, which allows the R_e_ values to change between <1 (0.193) to >5. A normal prior with M=48.7 and S=15 (corresponding to a 95% interval from 24.0 to 73.4) was used for the rate of becoming uninfectious. These values are expressed as units per year and reflect the inverse of the time of infectiousness (5.3-19 days, mean 7.5) according to the serial interval estimated by Li *et al*. [19]. Sampling probability (ρ) was estimated assuming a prior Beta (alpha=1.0 and beta=999), corresponding to a minority of the sampled cases (between 10^−5^ to 10^−3^). The origin of the epidemic was estimated using a normal prior with M=0.1 and S=0.05 in units per year.

The MCMC analyses were run for 100 million generations and sampled every 10,000 steps.

Convergence was assessed on the basis of ESS values (ESS >200). Uncertainty in the estimates was indicated by 95% highest posterior density (95%HPD) intervals.

The mean growth rate was calculated on the basis of the birth and recovery rates (r=λ-δ), and the doubling time was estimated by the equation: doubling time=ln(2)/r [20].

## 3. Results

### 3.1 Phylogenetic analysis of the whole dataset

No recombination events were observed in the entire dataset according to analyses with RDP5 software.

Phylogenetic analysis by maximum likelihood showed that the Italian sequences were included in a single SARS-CoV-2 clade (the old Nextstrain A2 clade) with the exception of three sequences: two from Chinese patients visiting Italy at the end of January 2020 after being infected in Wuhan and one characterized by us from an Italian subject, living in Padua, sampled in March 2020, not reporting any recent trip outside Italy or contacts with subjects affected by COVID-19 (pp=0.99) (Figure 1, clade 19A).

**Figure 1.**
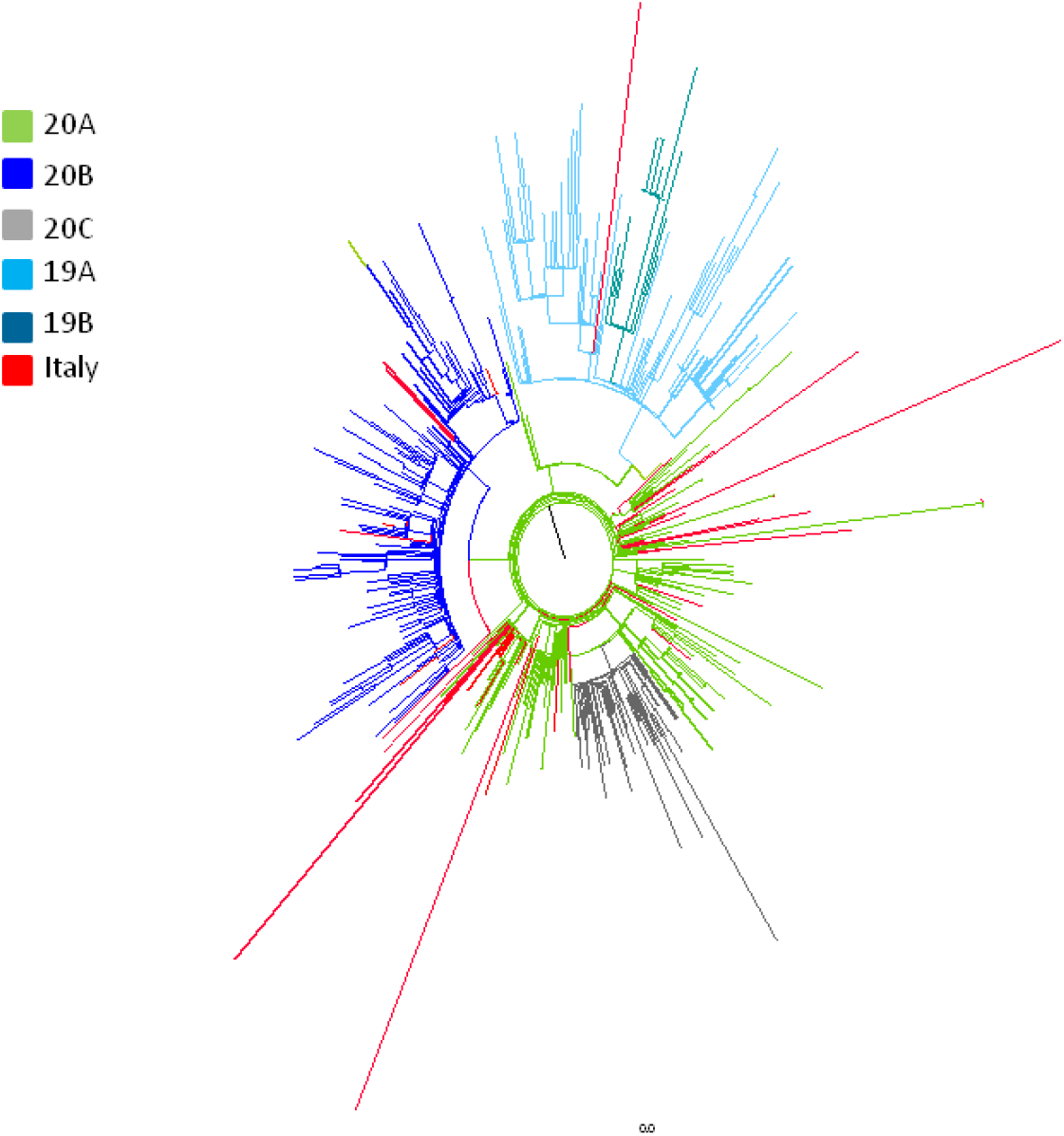
Maximum likelihood tree of the full dataset including 588 SARS-CoV-2 genomes. Nextstrain classification is indicated by colours as reported in the legend. Italian strains are highlighted in red.

Recently, new nomenclature systems have been proposed for the SARS-CoV-2 clades. The new lineage assignment of 62 Italian isolates is reported on Table 1 with the correspondence to other naming systems (old and new Nextstrain). All of our isolates belonged to the lineage B.1, only one isolate was classified as lineage B.

**Table 1.** Pangolin lineage classification of 62 Italian strains included in the study.

| Lineage (Pangolin) | Total | % | From | Nextstrain new | Nextstrain old |
| --- | --- | --- | --- | --- | --- |
| B | 1 | 1.6 | PD (1) | 19A | nd |
| B.1 | 47 | 75.8 | MI (15), PS(7), AN (1), MC (1) PD (8), BG (1), CR (3), SI (3), AR (3),<br>GR (1), BS (4) | 20A, nd | A2a |
| B.1.1 | 11 | 17.7 | MI (4), PD (1), SI (4), GR (1), AR (1) | 20B | A2a |
| B.1.34 | 1 | 1.6 | MI (1) | nd | A2a |
| B.1.5 | 2 | 3.2 | MI (1), BG (1) | 20A | A2a |

PD: Padua, MI: Milan, PS: Pesaro, AN: Ancona, MC: Macerata, BG: Bergamo, CR: Cremona, SI: Siena, AR: Arezzo, GR: Grosseto, BS: Brescia, nd: not determined.

### 3.2 Genetic distances analysis

The overall mean p-distance between all the Italian isolates was 2.3 (SE:0.3) s/10,000 nts, corresponding to a mean of 6.4 (SE: 0.8) substitutions per genome. The non-synonymous distance (dN) was 2.0 (SE: 0.4) non-syn s/10,000 non-syn nts while the overall synonymous mean distance (dS) was equal to 2.4 (SE: .05) syn s/10000 syn nts (dN/dS=0.83). A higher heterogeneity was observed through months as, stratifying the genetic distances on the basis of the sampling time, we observed a higher heterogeneity among the strains isolated in February (n=19) compared to those collected in March (n=96) or April (n=21) (Table 2).

**Table 2.** Mean genetic divergence within and between Italian strains according to the sampling time (substitutions per 10,000 sites).

| Time | Within |  |  |  | Time | Between |  |  |  |
| --- | --- | --- | --- | --- | --- | --- | --- | --- | --- |
|  | P distance (SE) | nucleotide (SE) | dS (SE) | dN (SE) |  | P distance (SE) | nucleotide (SE) | dS (SE) | dN (SE) |
| February | 3.8 (0.6) | 9.6 (1.5) | 3.5 (1.1) | 3.8 (0.6) | February vs March | 3.1 (0.4) | 8.1 (1.3) | 2.9 (0.8) | 2.8 (0.4) |
| March | 1.9 (0.3) | 5.4 (0.8) | 2.2 (0.5) | 1.5 (0.4) | March vs April | 2.3 (0.3) | 6.6 (0.8) | 2.1 (0.6) | 2.0 (0.5) |
| April | 2.4 (0.3) | 6.8 (0.9) | 1.7 (0.8) | 2.1 (0.5) | February vs April | 3.7 (0.5) | 10 (1.5) | 2.7 (0.8) | 3.5 (0.6) |

SE: Standard error, dS: synonymous distance, dN: non-synonymous distance.

### 3.3 Differences in Amino Acids

Considering only the non-synonymous mutations and comparing the Italian genomes with the common ancestor (China), there were 159 amino acid substitutions affecting different viral genes, (112 in ORF 1a/1b, 19 in S, 12 in ORF 3a, 4 in M, 3 in ORF7a, 6 in N, and one each in Orf7b, 8 and 10) of which only 15 (9.4%) were observed in 2 or more isolates, as summarized in Table 3. No aminoacid changes were observed in the E gene. The previously described substitution D614G in the Spike protein was present in all the isolates belonging to the lineage B.1 and in the strain from Padua belonging to lineage B.

**Table 3.** SARS-CoV2 mutations identified in Italian strains.

| Genome<br>region | Mutation | n/total | Percentage<br>(%) |
| --- | --- | --- | --- |
| ORF 1ab | S443F | 2/135 | 1.5 |
|  | H3076Y | 2/135 | 1.5 |
|  | L3606F | 3/131 | 2.3 |
|  | P4715L | 133/136 | 97.8 |
|  | E5689D | 2/135 | 1.5 |
|  | R5919K | 2/123 | 1.6 |
| S | A570D | 2/129 | 1.6 |
|  | D614G | 128/130 | 98.5 |
|  | G1046V* | 3/134 | 2.2 |
| ORF 3a | G251V | 3/134 | 2.2 |
| M | D3G | 21/133 | 15.8 |
| ORF 7a | G70C | 2/134 | 1.5 |
| N | R203K-G204R | 52/133 | 39.1 |
|  | V246I | 3/136 | 2.2 |

\* mutation under significant selective pressure

Considering the Italian isolates, only 1 site resulted under significant selecting pressure by three different methods (MEME, FEL, FUBAR): site 1,046 in the S gene that was present in three isolates from Padua. This G1046V mutation is located in the S2 subunit, between heptad repeat 1 and 2. Mutations R203K-G204R in N gene were always simultaneously detected. It appears that these mutations discontinue a serine-arginine (S-R) dipeptide by introducing a lysine in-between them, having impacts on structure and function in the mutated N protein.

Fifty two sequences in our dataset carried these mutations, particularly 11 of the 59 whole genome newly characterized; six of these were from Tuscany, four from Milan and one from Padua.

### 3.4 Time reconstruction of the SARS-CoV-2 Italian lineage B.1 phylogeny

Root-to-tip regression analysis of the temporal signal from the Italian B.1 subset revealed a weak association between genetic distances and sampling days (a correlation coefficient of 0.31 and a coefficient of determination (R^2^) of 9.9 × 10^−2^).

Comparison by BF test of the marginal likelihoods obtained by path sampling (PS) and stepping stone sampling (SS) of the strict vs relaxed molecular clock (uncorrelated log-normal) showed that the second performed better than the former (strict vs. relaxed molecular clock BF(PS)=-71.9 and BF(SS)=-71.4 for relaxed clock). Comparison of the different demographic models showed that the BSP and the exponential growth models best fitted the data (BSP vs. constant population size BF(PS)= 27.9 and BF(SS)= 30.2 for BSP; constant population size vs. exponential growth BF(PS)= 7.3 and BF(SS)= 8.6) (Table S3).

The mean tMRCA of the tree root (Figure 2) was estimated at 107 days before present (BP) (95%HPD: 91.2-113.1), corresponding to January 11 2020 (from January 5 to January 27). The tMRCA of the subclade including all the Italian sequences was estimated to be 92.4 (95%HPD: 76.6-95) days BP, corresponding to January 25 (between January 23 and February 10).

**Figure 2.**
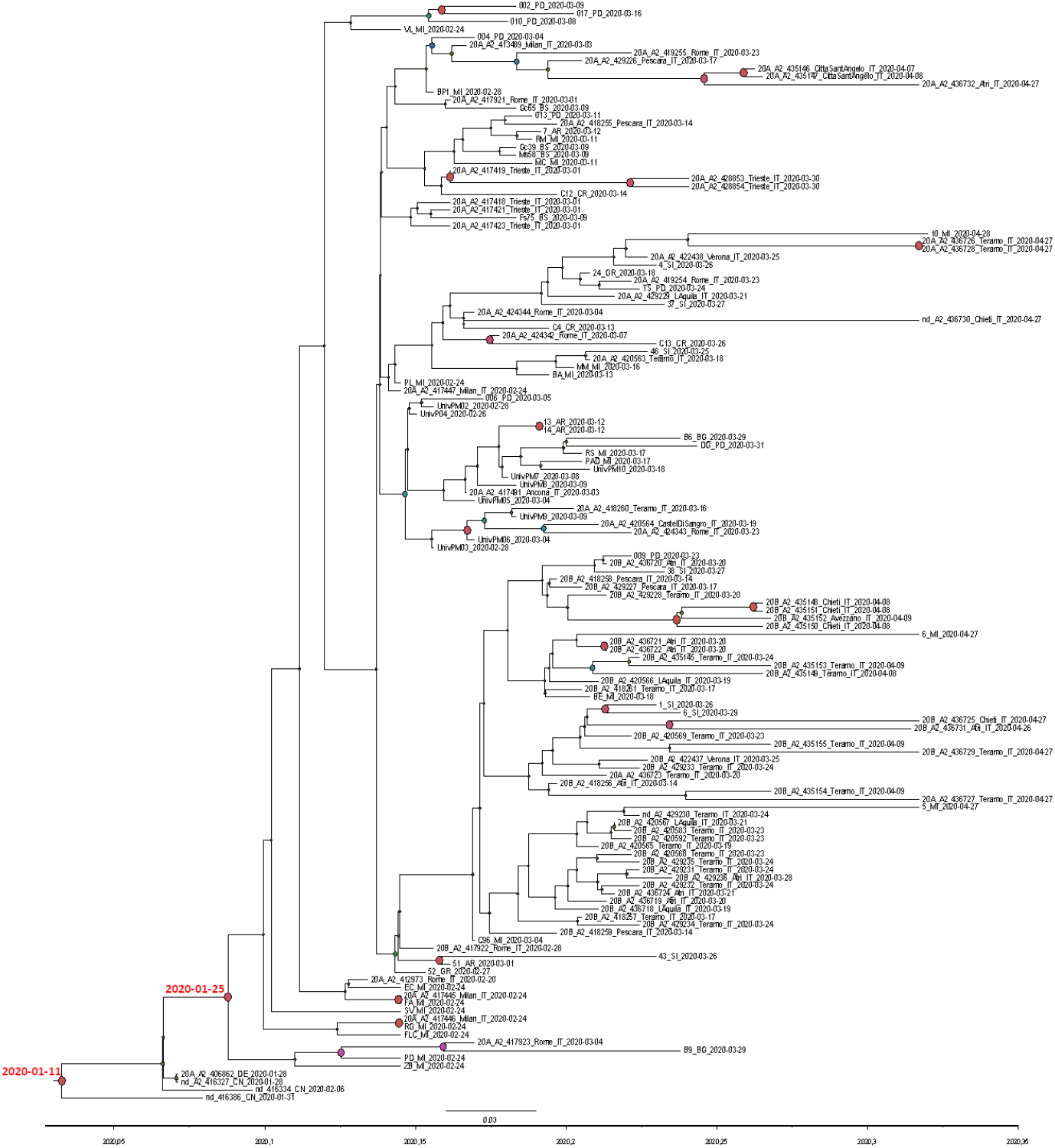
SARS-CoV-2 tree of 136 Italian strains plus one German and three Chinese isolates from Shanghai, showing statistically significant support for clades along the branches (posterior probability > 0.7).Large red and purple circles indicated highest posterior probability. Calendar dates of the tree root and the Italian clade were showed in red.

The Bayesian tree of the Italian sequences showed 15 small significant subclades including two to ten isolates (Figure 2).

### 3.5 Phylodynamic analysis of the Italian dataset

The Bayesian skyline plot of the Italian isolates showed an increase in the number of infections in the period between 23 February and mid-March 2020, with a rapid exponential growth between March 4 and 16 when it reached a plateau continuing until the last sampling time (Figure 3).

**Figure 3.**
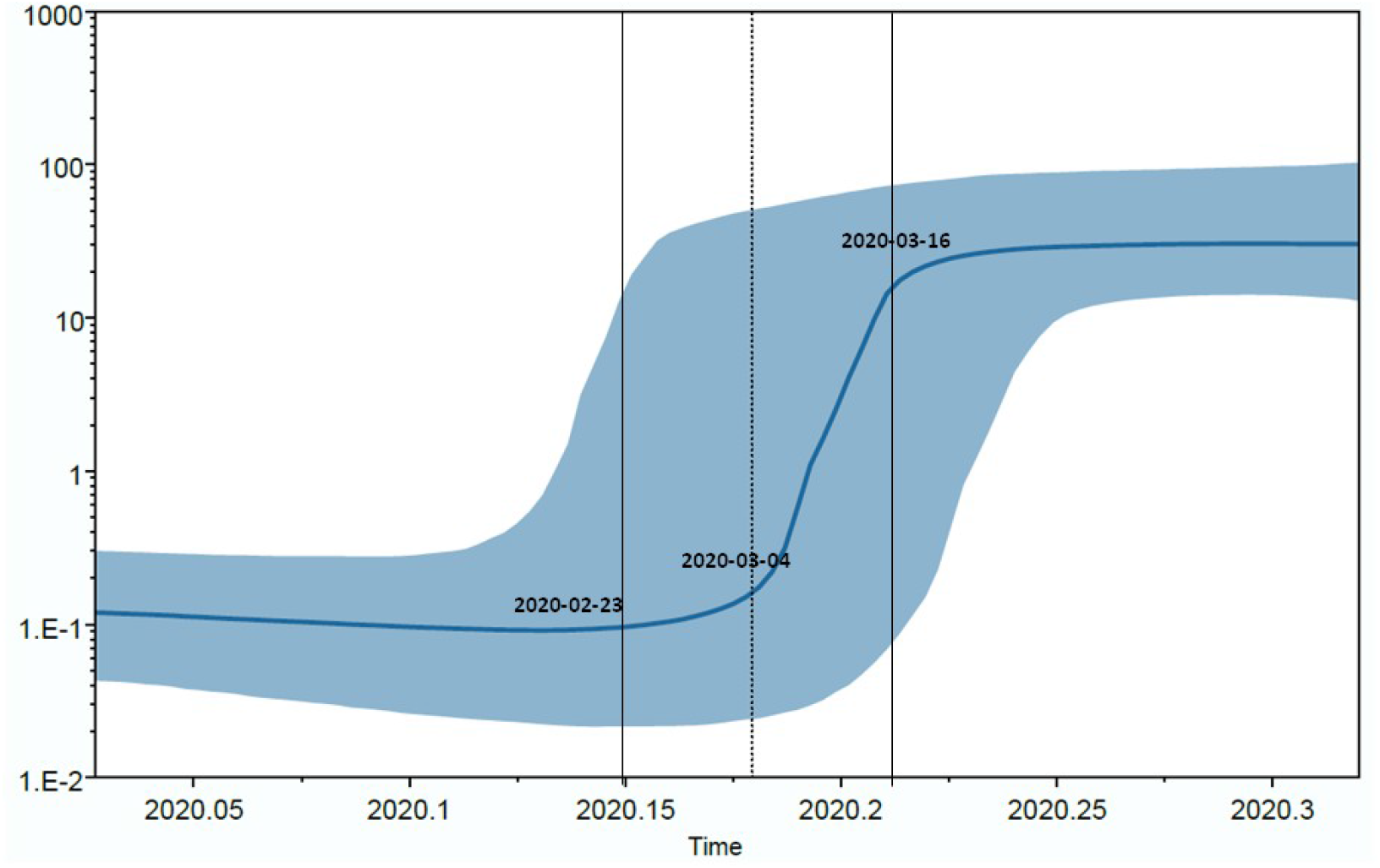
Bayesian Skyline plot of the SARS-CoV-2 outbreak. The Y axis indicates effective population size (Ne) and the X axis shows the time in fraction of years. The thick solid line represents the median value of the estimates, and the grey area the 95% HPD.

The Bayesian birth-death skyline plot of the R_e_ estimates with 95%HPD with a single R group (corresponding to R_0_) estimated a mean value of 2.25 (1.5-3.1). Figure 4 (panels a and b) shows the changes of R_e_ since the origin of the epidemic and suggests that R_e_ was higher than 1 since the early days (mean initial R_e_=1.4, 95%HPD: 0.08-2.9). The curve started to grow in early February and peaked to a mean value of 2.3 (95%HPD: 1.5-3.5) in the first half of March, and has since remained at this value. The curve obtained with three R_e_ groups showed a slight decrease at mid-March (Figure 4, panel b).

**Figure 4.**
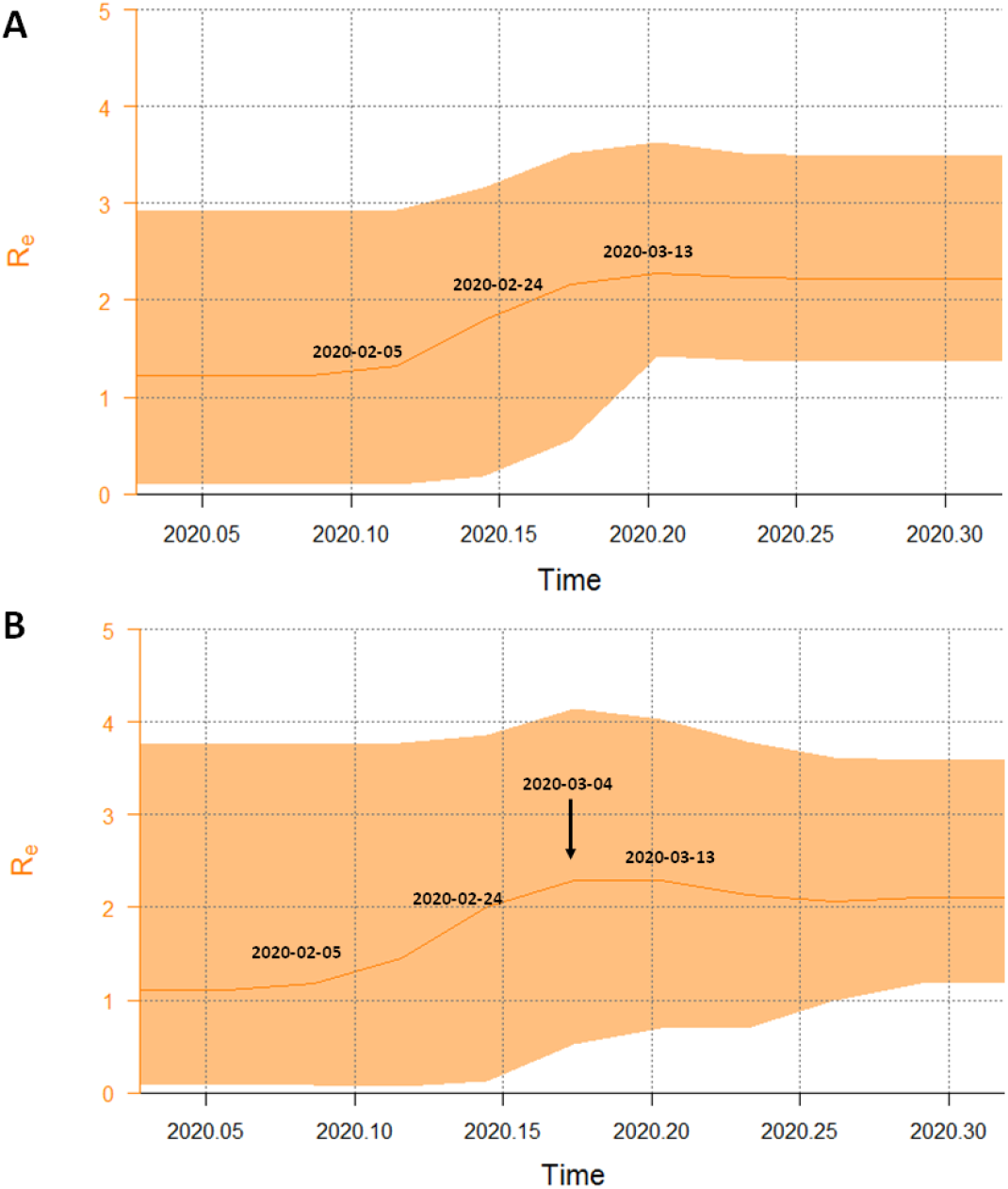
Part A: Birth-death skyline plot of the SARS-CoV-2 outbreak allowing one R_e_ intervals. Part B: Birth-death skyline plot of the SARS-CoV-2 outbreak allowing three R_e_ intervals. The curves and the orange areas show the mean R_e_ values and their 95% confidence intervals. The Y and X axes indicate R values and time in years, respectively.

The origin of the epidemic was estimated at a mean 80.3 days BP (credibility interval: 60-109), corresponding to February 7 (between January 9 and February 27). The recovery rate was estimated about 7.26 days (CI 4.7-16.0 days), and the transmission rate (λ) increased from 71.7 to 115.96 in units per year (corresponding to a growth rate of 0.06 and 0.18 year^-1^). On the basis of these data, the doubling time decreased from 5.1 days to 3.1 days in the period between early February and mid-March.

## 4. Discussion

Molecular tracing of SARS-CoV-2 coupled with advanced Bayesian and Maximum likelihood phylogenetic analysis provide detailed information about the epidemiology and evolution of emerging infections and helps to improve our understanding on the mechanisms of spreading of the epidemic.

In a previous study [1], we characterized the viral sequences obtained from the first three patients coming from the Codogno area who were hospitalized at the very beginning of the epidemic in Italy. The Codogno strains correlated with an isolate from an outbreak occurred in Bavaria around January 20 [4]. The present analysis shows that all but one of 62 SARS-CoV-2 sequences obtained from February 22 to the end of April in different Northern and Central Italian areas belong to a single clade, corresponding to the Pangolin lineage B.1, the old Nextstrain subclade A2a and the new Nextstrain clades 20A and 20B (https://nextstrain.org/blog/2020-06-02-SARSCoV2-clade-naming) [12], [1]. About 1 out of 4 isolates were classified in different clusters, always included in the main B.1 lineage (such as B.1.1 and B.1.5), most on a temporal basis, being these clusters more represented among the genomes sampled in the second half of March and April (9/14, 64%), while B.1 lineage was more represented in the genomes obtained in February and first half of March (33/47, 70.2%).

This observation was also confirmed by other Italian studies [3], [1]. The same clade is now the most widespread in the world and includes most of the published genomes [5]. The genetic distances among the Italian strains were relatively short, corresponding to an average of about 6.4 mutations per viral genome, even if single isolates may have a higher number of changes. After grouping the sequences according with the sampling months, while the within group mean genetic distances were higher in February compared to subsequent months, the genetic distance between different months increased with time. This observation confirms a continuous evolution of the viral genome (with the emergence of new divergent variants) mainly driven by genetic drift. No significant difference was observed between the non-synonymous and the synonymous substitutions (dn/ds=0.8), suggesting the absence of relevant selective forces driving the evolution of the viral genome. This observation is further confirmed by the analysis of site-specific selective pressure in the Italian strains, which only showed a single site under significant positive selection in the S protein (position 1,046) observed in three strains from Padua. Including in the phylogenetic tree 3 isolates from Shanghai and one from the first patient of the Bavarian cluster, being at the root of the B.1 lineage, the dated tree obtained suggests that SARS-CoV-2 entered Italy between late January and early February 2020. This timing matches with the first autochthonous European cluster of SARS-CoV-2 transmission in Bavaria (Germany), originated on January 20 [21], [4], [1] by the introduction of a strain carried by the index patient coming from Shanghai, where the virus had been circulating since January. The skyline plot analysis of the Italian clade shows an exponential increase of the effective number of infections from late February to mid-March, in excellent agreement with the known epidemiological data (https://www.epicentro.iss.it/coronavirus/sars-cov-2-dashboard). In particular, a very rapid growth of the epidemic was detected between the beginning of March and the middle of the same month, when the curve reaches a plateau up to the end of sampling (27 April). The mean value of R_0_ was estimated as 2.25 (1.5 to 3.1) in the entire period. A similar result was obtained by Stadler et al. on a smaller sample of 11 sequences mainly from patients with known travel history to Italy (https://virological.org/t/phylodynamic-analyses-based-on-11-genomes-from-the-italian-outbreak/426). The estimated basic reproduction number (*R*0) for SARS-CoV-2 has ranged mainly from 2 to 4, according to the different methods employed for the evaluation [22]. In Italy, values between 2.4 and 3.6 have been estimated in the early phase of COVID-19 epidemic before the control measures were taken [23], [24], [25]. Predictive mathematical models are fundamental to understand the dynamics of the epidemic, plan effective control strategies and verify the efficacy of those applied.

Using a birth-death skyline, we analysed the changes of R_e_ during the epidemic in Italy over the entire period. We observed that the R_e_ was >1 since the first decade of February, suggesting that the infection was circulating within the population before the first notified (hospitalized) COVID-19 cases. The R_e_ skyline plot reached a value of 2.3 in the first days of March, together with the rapid increase observed in the number of infections by BSP, and slightly decreased thereafter, in agreement with the official data on the course of the epidemic. Between February and March the estimated doubling time of the epidemic decreased from 5.1 to 3.1 days. This value was smaller than that obtained by us for the epidemic in China [26] and might be interpreted as a consequence of a delayed application of more stringent containment measures in Italy. In fact, a slight decrease of the R_e_ value was observed only after mid-March, when a more rigorous social distancing was enforced across the entire country. The persistence of a R_e_ value higher than one until April, in partial contrast with the epidemiological data (https://covstat.it/), could be due to the fact that our estimate was influenced by the circulation of the virus in the community, which is larger than the number of the officially registered clinical cases. It is well known that only a small minority of SARS-CoV-2 infections require hospitalization and that in Italy the number of cases of infection has widely exceeded the number of official reports. In a recent study, the prevalence of anti-SARS-CoV-2 antibodies in asymptomatic blood donors living in Milan was shown to increase from February to April, when the prevalence reached its maximum (about 7%) [27]. However, in Italy the numbers of active cases began to decrease only in the second half of April, when the present study had already been stopped. Further studies on extended data collection will be required to estimate the effects of the containment measures.

The only one genome characterised in our study not belonging to lineage B.1 was isolated in a 76-year-old man living in the province of Padua (Veneto), who survived to serious COVID-19 manifestations despite old age and the presence of several comorbidities. He denied any contact with infected subjects and did not travel abroad. This virus belongs to the same lineage (B) of the first 2 cases imported into Italy from the Hubei region, China, at the end of January 2020, before Italy suspended flights from China. The couple landed at the Milan airport and travelled to other locations in Northern and Central Italy before the onset of symptoms requiring hospitalization in Rome, but they had not travelled to Padua. Thus, the origin of such a strain remains unexplained and further investigations are underway to evaluate whether this strain may have played a role in causing an epidemic, at least locally. It would also be interesting to investigate whether the currently predominant strain was for some reasons more epidemic than the initial strain, or if the spread of the latter was limited by random factors.

In conclusion, our data show the importance of molecular and phylogenetic evolutionary reconstruction in the surveillance of emerging infections. Of note, it appears that the outbreak in Italy, which involved hundreds of thousands of people, is mainly attributable to a single introduction of the virus and its uncontrolled circulation for a period of about four weeks. These results reaffirm the strategic importance of continuous surveillance and timely tracing to define and rapidly implement effective containment measures for a possible second wave of the pandemic.

## Author Contributions

Conceptualization, A.L, G.Z, C.B., M.G. methodology, A.L, G.Z.; software, A.L, G.Z., A.B.; formal analysis, A.L, G.Z., S.R., A.B.; investigation, A.L., A.B., N.C., I.V., F.D., S.M., F.C.; writing—original draft preparation, A.L, G.Z, M.G.; writing—review and editing, A.L, G.Z, M.G., A.B., C.B; visualization, all authors; supervision, all authors; project administration, G.Z, C.B., M.G.; funding acquisition, G.Z, M.G. All authors have read and agreed to the published version of the manuscript.

## Data Availability

All new sequences will be available on GISAID.

## Acknowledgments

We acknowledge the authors and the originating and submitting laboratories of the GISAID sequences. The research was conducted under a cooperative agreement between Università degli Studi di Milano - Medicina del Lavoro e Clinica delle Malattie Infettive del Dipartimento di Scienze Biomediche e Cliniche “Luigi Sacco”, Intesa Sanpaolo and Intesa Sanpaolo Innovation Center.

## Contributor Information

SCIRE collaborative Group

F Alessandrini1, M Andreoni2, G Antonelli3, S Babudieri4, P Bagnarelli5, S Bonora6, B Bruzzone7, G Brindicci8, P Carrer9, F Ceccherini4, M Codeluppi10, A Coluccello11, N Coppola12, MG Cusi13,14, C Della Ventura9, L Di Sante5, S Di Giambenedetto15, G Di Perri6, V Fiore4, S Fiorentini16, D Francisci17, C Gandolfo13, C Gervasoni18, V Ghisetti6, R Greco19, G Guarona20, M Iannetta2, L Li Puma21, V. Malagnino2, G Mancuso 22, C Mastroianni3, I Menozzi23, E Milano8, A Miola21, M Morganti23, L Monno8, G Noberasco20, G Nunnari22, V Onofri1, A Orsi20, MG Pierfranceschi11, S Pongolini23, D Ripamonti24, S Rubino4, L Ruggerone21, D Russignaga25, C Sagnelli12, M Sanguinetti15, L Sarmati2, L Sasset26, E Scaltriti23, R Schiavo10, M Schioppa27, S Testa11, C Turchi1, O Turriziani3, E Venanzi Rollo22, S Zanussi28, A Zoncada11

1. Section of Legal Medicine, Polytechnic University of Marche, Ancona, Italy
2. University of Rome Tor Vergata, Rome, Italy
3. Laboratory of Virology, Department of Public Health and Molecular Medicine, Sapienza University, Rome, Italy.
4. Department of Medical, Surgical, and Experimental Sciences, University of Sassari, Viale San Pietro 43, 07100 Sassari, Italy
5. Department of Biomedical Sciences and Public Health, Virology and Legal Medicine Laboratories, Polytechnic University of Marche, Ancona, Italy
6. Clinic of Infectious Diseases, Amedeo di Savoia Hospital, University of Torino
7. Hygiene Unit, IRCCS AOU San Martino-IST, Genova, Italy
8. University Hospital Consortium of the Polyclinic of Bari, Bari, Italy
9. Department of Biomedical and Clinical Sciences “L. Sacco” University of Milan, IT-20157 Milan, Italy
10. Health Unit of Piacenza, Piacenza, Italy
11. Unit of Infectious Diseases, Territorial Social Health Company of Cremona, Cremona, Italy
12. Department of Mental Health and Public Medicine, Section of Infectious Diseases, University of Campania Luigi Vanvitelli, Naples, Italy
13. Department of Medical Biotechnologies, University of Siena, Siena, Italy
14. Microbiology and Virology Unit, Siena University Hospital, University of Siena, Italy
15. Institute of Clinical Infectious Diseases, Catholic University of Sacred Heart, Rome, Italy
16. Laboratory of Microbiology and Virology, Territorial social health companySpedali Civili, Department of Molecular and Translational Medicine, University of Brescia, Italy
17. Infectious Diseases Clinic, Department of Medicine, University of Perugia, Perugia, Italy
18. ASST Fatebenefratelli Sacco, III Division of Infectious Diseases, University of Milan
19. Microbiology Unit, Clinical Pathology Complex Operative Unit, Sant’Anna e San Sebastiano Hospital, Caserta, Italy
20. Department of Health Sciences (DISSAL), University of Genoa, Genoa, Italy
21. Intesa Sanpaolo Innovation Center, Turin, Italy
22. Complex Operating Unit, Infectious Diseases, Policlinico Gaetano Martino, University of Messina, Italy
23. Risk Analysis and Genomic Epidemiology Unit, Experimental Prophylactic Zoo Institute of Lombardy and Emilia Romagna, Parma, Italy
24. Infectious Diseases Unit, Territorial Social Health Agency of Bergamo, Papa Giovanni XXIII Hospital, Bergamo
25. Prevention and Protection Service, INTESA S.p.A., Turin, Italy
26. Infectious and Tropical Diseases, University of Padova-Hospital, Padova, Italy
27. Simple departmental operating unit of Genetics and molecular biology, AORN S. Anna e S. Sebastiano, Caserta, Italy
28. COS of Medical Oncology and Immunocorrelated Tumors, Oncological Reference Center, Aviano, Italy

